# Deep Learning and Radiomics Models for Predicting 90-Day Outcomes in Ischemic Stroke: A Systematic Review and Meta-Analysis

**DOI:** 10.64898/2026.10.13.26365396

**Authors:** Iman Kiani, Kimia Darmiani, Parya Valizade, Payam Jannatdoust, Sanaz Bordbar, Amir Hossein Majidzade Moqadam, Bardia Khosravi

**Affiliations:** Students’ Scientific Research Center, Tehran University of Medical Sciences, Tehran, Iran; Department of Neuroradiology, Rockefeller Neuroscience Institute, West Virginia University, Morgantown, WV 26505; School of Medicine, Tehran University of Medical Sciences, Tehran, Iran; Joint Department of Medical Imaging, University Health Network, Toronto, Ontario, Canada; Assistant Professor, Department of Radiology, Mayo Clinic, 20011st St SW, Rochester, MN 55905

**Keywords:** Stroke, 90-day mRS, deep learning, radiomics

## Abstract

**Background:** Accurate prediction of 90-day outcomes following ischemic stroke is crucial for optimizing clinical strategies. This study aimed to systematically review and meta-analyze studies using deep learning (DL) and hand-crafted radiomics (HCR) models for predicting 90-day modified Rankin Scale (mRS) outcomes after ischemic stroke.

**Methods:** We followed the PRISMA guidelines for systematic reviews. Four databases (PubMed, Embase, Web of Science, and Cochrane Library) were searched comprehensively for studies published up to September 2025. Inclusion criteria encompassed original research focusing on ischemic stroke patients with mRS at 90 days as the primary outcome and predictive models utilizing imaging biomarkers from CT or MRI. Quality assessment of the included studies was conducted using the PROBAST and and METhodological RadiomICs Score (METRICS) tools.

**Results:** Seventeen studies were included in the quantitative meta-analysis. Models demonstrated good prognostic performance for 90-day outcomes, with pooled AUCs of 0.85 (95% CI: 0.80–0.86) for internal and 0.74 (95% CI: 0.67–0.89) for external validation sets, with no significant difference (p=0.42). Deep learning and hand-crafted radiomics models showed comparable diagnostic accuracy in the primary analysis (p=0.667); a difference in sensitivity favouring hand-crafted radiomics emerged only in a post-hoc analysis after exclusion of one outlier study. Models incorporating clinical variables significantly outperformed imaging-only models (p<0.01). CT- and MRI-based models showed similar performance. Treatment-specific analyses revealed higher sensitivity for IVT-based models and higher specificity for combined MT+IVT models.

**Conclusions:** DL and HCR models demonstrate robust potential for predicting 90-day outcomes in ischemic stroke. Future research should focus on standardizing model evaluation metrics and incorporating explainable AI techniques to improve clinical applicability.

## Introduction

Stroke is regarded as the second cause of death and a leading cause of disability in the world [1]. Patients from stroke expereince function loss and disabilities that significantly impact their quality of life [2, 3]. Studies have shown up to one-third of patients with ischemic strokes who have undergone appropriate treatment still exhibit poor functional outcomes at 90 days [4, 5], as measured by modified Rankin Scale (mRS) score greater than two. Therefore, it is of high importance to predict the prognosis and long-term disabilities of patients with ischemic strokes to optimize strategies for rehabilitation and clinical management [6]. However, this prediction is challenging due to multiple factors that affect it [7].

Various methods have been used to predict the prognosis of patients with stroke. MRI findings, such as infarct volume, have been applied for the prediction of functional outcomes [8]. Besides imaging biomarkers, clinical data such as comorbidities, neurological deficits, and age can be used for predicting stroke outcomes [9]. Additionally, traditional scoring systems, such as the Totaled Health Risks in Vascular Events score (THRIVE), have historically been used to predict stroke outcomes [10].

However, in recent years, artificial intelligence (AI) methods have been employed to improve the predictive power of models. Deep learning (DL) based methods such as convolutional neural networks (CNN) and transformer based can comprehend medical images and combine the findings with clinical data. They can extract and identify clinical features for predicting long-term outcomes without prior feature engineering [11]. Additionally, hand-crafted radiomic (HCR) features have proved to hold valuable information [12]. HCR involves rule-based extraction of high-dimensional quantitative features from medical imaging, such as CT and MRI. These features capture information about shape, texture, intensity, and spatial relationships within the imaging data [13], which in the field of stroke, have been linked to biological processes such as the clot histopathology [14].

Given the increasing integration of neuroimaging with computational tools, assessment of the models on predicting stroke outcome has become a critical priority. While both HCR and DL models have demonstrated potential in this domain, they differ substantially in methodology, interpretability, and data requirements. Additionally, the choice of imaging modality, whether CT or MRI, may influence predictive performance. Therefore, this study aims to conduct a systematic review and meta-analysis of existing literature applying HCR and DL-based models to predict 90-day functional outcomes in stroke patients based on CT and MRI.

## Methods

This review complies with the Preferred Reporting Items for Systematic Reviews and Meta-Analyses (PRISMA). The protocol for this study was pre-registered at the International Prospective Register of Systematic Reviews (PROSPERO) with the ID CRD42024548342. Regarding ethical approval, given that this manuscript constitutes a review of previously published literature, it did not require an IRB approval number.

Two independent reviewers initially developed the search syntax through the use of both EMTREE and MeSH terms. When initial consensus was not reached, a third reviewer was consulted. The search was executed across four databases: PubMed, Web of Science, Embase, and Cochrane Library on September 15, 2025, as detailed in Table E1. A thorough examination of reference lists was conducted as well. Initial screening of records for duplicates was conducted using EndNote software, followed by manual clarification.

### Inclusion and exclusion criteria

For this review, we included studies that focused on patients with ischemic stroke and reported functional outcomes using the mRS at 90 days. Eligible studies were required to use CT or MRI for outcome prediction and apply either HCR or DL techniques to the imaging data. Studies focused on hemorrhagic stroke or other non-ischemic etiologies were excluded. We also excluded review articles, case reports, commentaries, and editorials.

### Screening and extraction

Two reviewers independently screened the search results, and irrelevant studies were removed. For quality assessment, the same two reviewers independently assessed bias risk using the Prediction model Risk of Bias Assessment Tool (PROBAST) and METhodological RadiomICs Score (METRICS) tool. Disagreements were adjudicated by a third reviewer as the final decision-maker. In addition, the two reviewers carried out data extraction independently. For cross-checking and verification, the extracted data were entered into an Excel spreadsheet. The third reviewer was consulted for unresolved issues.

### Data Synthesis

Following the completion of screening and data extraction, studies were selected for inclusion in a random-effects diagnostic test accuracy (DTA) meta-analysis based on the availability of these diagnostic metrics: true positives (TP), true negatives (TN), false positives (FP), and false negatives (FN). When multiple models were presented in a single study, the “primary” or best-performing model was chosen for the general meta-analysis, as indicated by the study’s accuracy metrics. “Secondary” models were considered for inclusion in subgroup analyses when relevant to specific hypotheses or contexts.

Cross-validated training cohorts were considered as internal validation sets. Subgroup analyses were conducted to compare the results from these sets to account for potential variations in performance between internal and external validation datasets. This was done to address common concerns such as overfitting and data mismatch. We hypothesized that performance differences would emerge based on whether models used DL algorithms for feature extraction or relied on HCR. Furthermore, we explored the potential influence of incorporating clinical variables alongside imaging features on diagnostic performance. We also hypothesized that the choice of pre-treatment imaging modality (CT vs. MRI) and the specific treatment type (mechanical thrombectomy (MT) alone, intravenous thrombolysis (IVT), or combined MT+ IVT) could influence the diagnostic accuracy of the predictive models. SROC curves were generated when at least three models were available within each subgroup.

The bivariate model proposed by Reitsma et al. was applied to the DTA meta-analysis [15]. Summary Receiver Operating Characteristic (SROC) curves were derived from the bivariate meta-analysis, with study-specific estimates weighted according to their contribution within a random-effects univariate Diagnostic Odds Ratio (DOR) framework. The Area Under the SROC Curve (AUC) and its confidence intervals were estimated for each subgroup using a bootstrap sampling approach (n=2000) within the bivariate model framework [16].

Heterogeneity among studies was assessed using the I² statistic, in accordance with the Holling et al.’s [17] method, with values exceeding 25% suggesting significant heterogeneity [18]. Sensitivity analyses were performed based on a DOR univariate model to identify and address any outlier studies, which were then re-analyzed to ensure result consistency. Furthermore, publication bias was evaluated using a modified version of Egger’s regression test adapted for DTA meta-analyses, analyzing funnel plot asymmetry based on 2000 bootstrap samples, following recommendations from Noma et al. [19].

All analyses were conducted with R software (version 4.2.1, R Foundation for Statistical Computing, Vienna, Austria), employing “MVPBT” [20], “Mada”, “dmetatools” [16], “meta” [21], and “Metafor” [22] packages.

## Results

### Overview

The PRISMA flowchart is shown in Figure 1. Initial screening identified 924 studies. After removing duplicates, 673 studies were screened, from which 46 studies were assessed for eligibility. Out of these, seventeen studies were included in the review [23–39]. The study characteristics are shown in Table 1. The studies were conducted across multiple countries, with the majority originating from China (n=9), followed by the USA (n=3). Regarding study settings, 10 studies were single-center while 7 utilized multi-center data. The training sample sizes ranged considerably from 36 to 1,613 patients, with validation cohorts ranging from 30 to 993 patients. In terms of imaging modalities, MRI was the most frequently used (n=10), followed by CT-based imaging (n=6), and one study utilized combined CT and intraoperative monitoring. Regarding analytical approaches, 9 studies employed deep learning methods while 8 utilized hand-crafted features. Model performance, as measured by AUC, ranged from 0.73 to 0.96, with most studies reporting AUCs between 0.80 and 0.92, indicating generally good discriminative ability for predicting 90-day functional outcomes in ischemic stroke patients.

**Figure 1.**
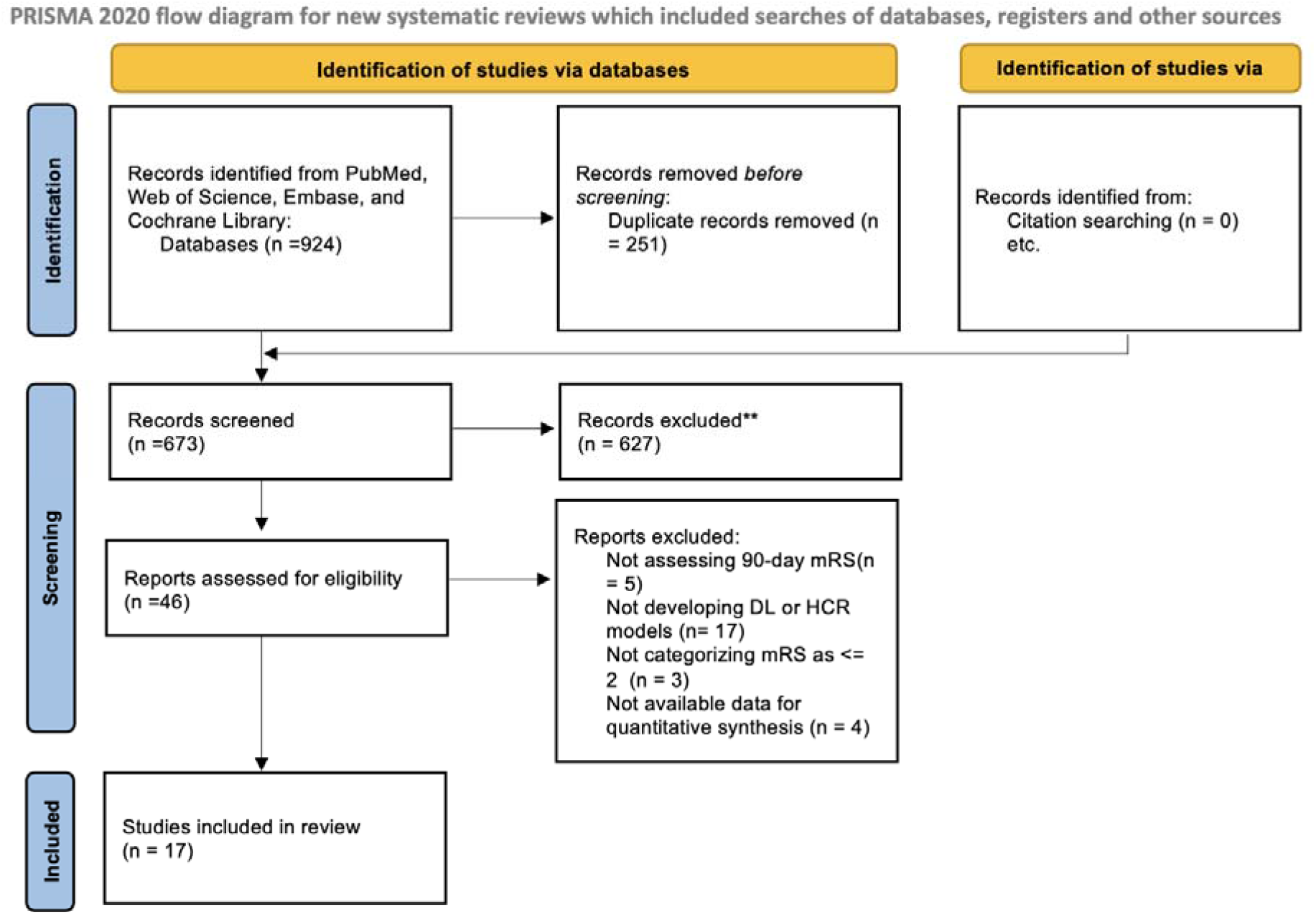
PRISMA flowchart

**Table 1.**
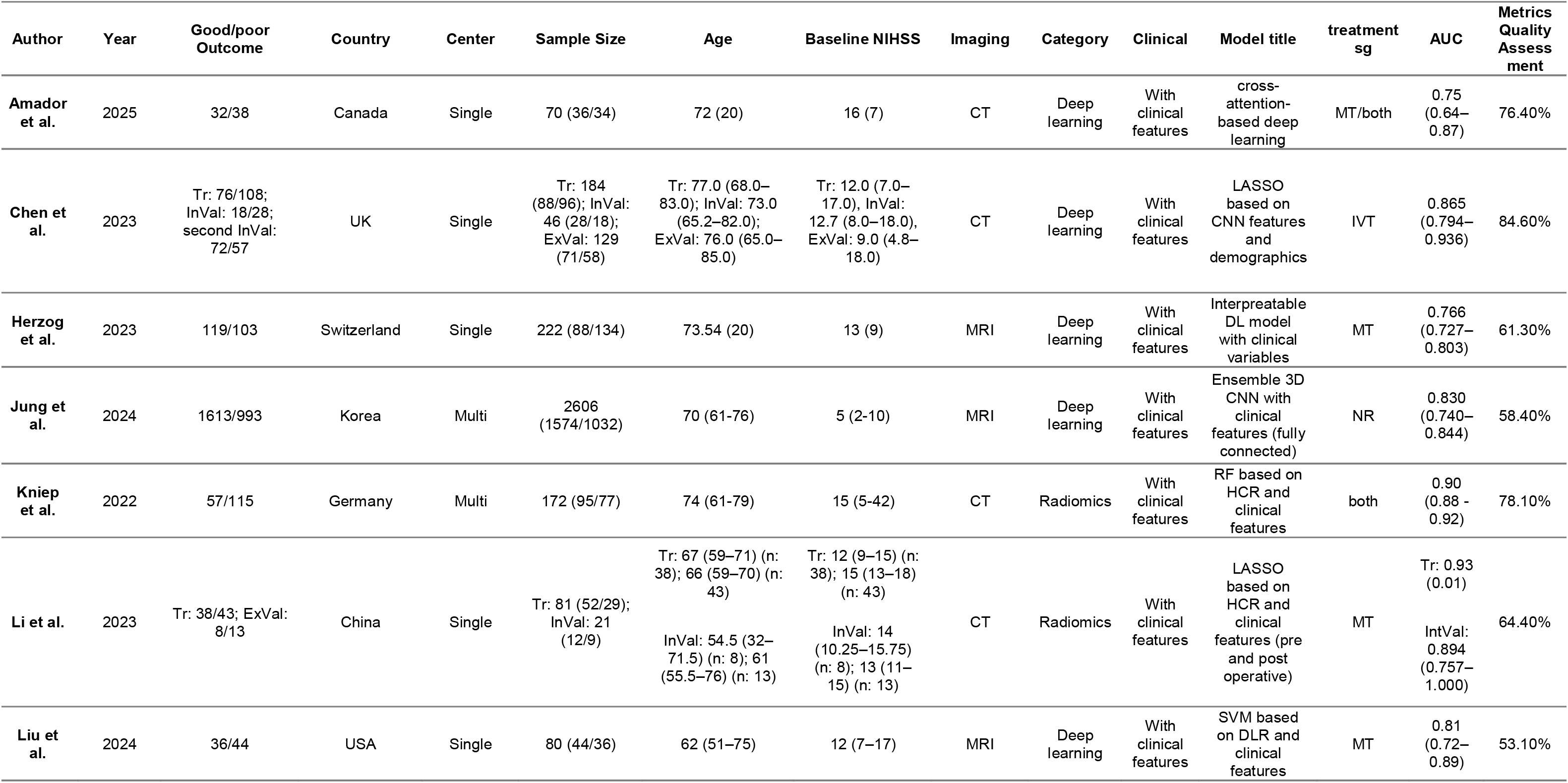
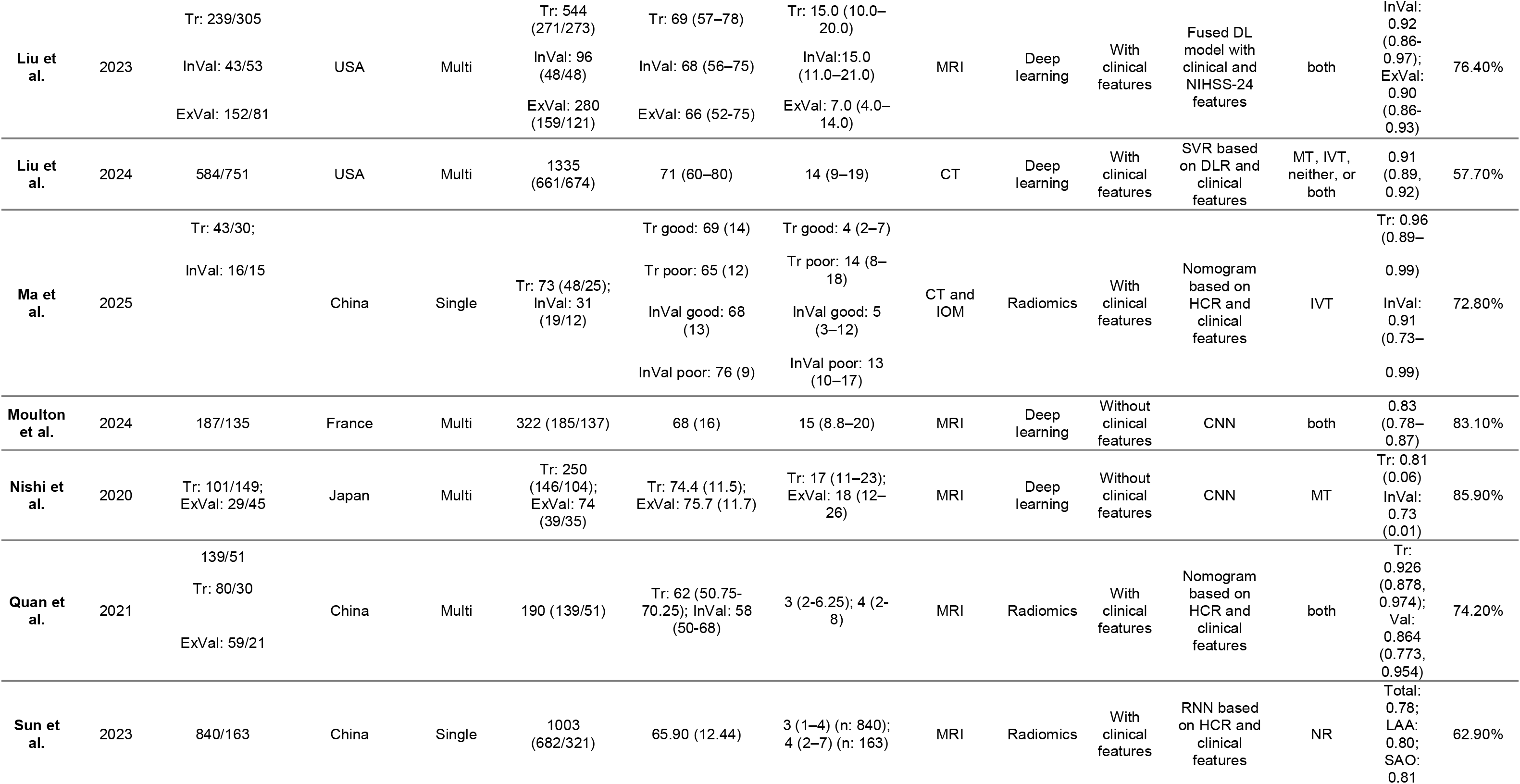
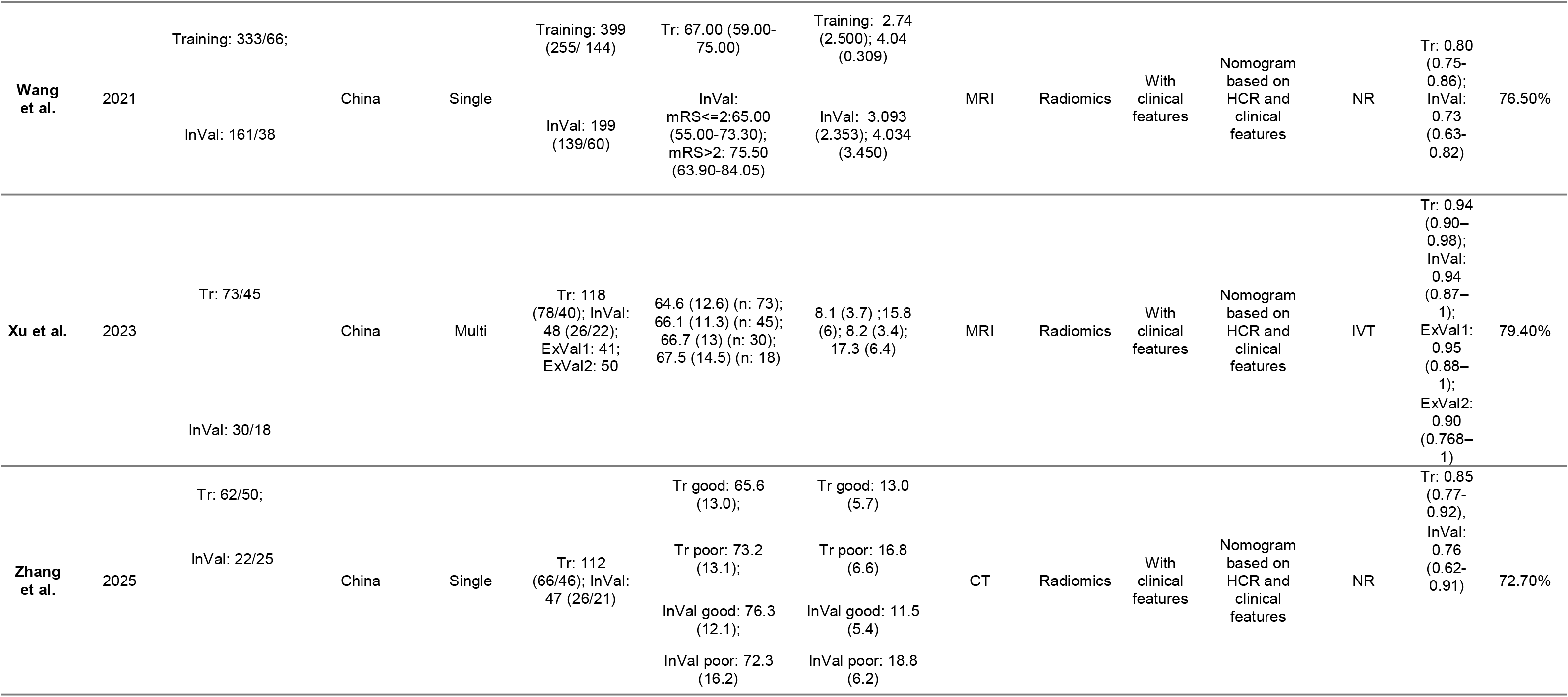
Summary of study characteristics.

METRICS results are also presented in Table 1. Overall, while the studies demonstrated adequate quality in the use of HCR and DL models to predict stroke outcomes, they often suffered from some limitations that affect their methodological quality. Most notably, few of the studies adhered to established radiomics or AI reporting checklists, and external validation was frequently absent. Calibration analysis, robustness testing (such as repeat-scan reliability or sensitivity analyses), and uncertainty quantification were generally lacking, undermining the reliability of the models in real-world settings. Public availability of data, code, and trained models was rare, limiting reproducibility and external benchmarking. Future work would benefit from stricter adherence to reporting standards, larger and more diverse multicenter datasets, open science practices, and a stronger focus on model generalizability and interpretability.

### Overall meta-analysis of models for 90-day prognosis in ischemic stroke: comparison of internal vs external validation sets

Among the reviewed studies, sufficient data were available from 17 studies to calculate the diagnostic accuracy metrics (TP, TN, FP, and FN) for their best predictive models. Studies lacking this data were excluded from further analysis. A qualitative synthesis of studies that were not eligible for inclusion in the meta-analysis is provided in the Supplementary Material.

Figure 2.A presents the SROC curves generated from the bivariate random-effects DTA meta-analysis of the best models of the included studies, stratified by internal and external validation datasets. The AUC was calculated as 0.85 (95% CI: 0.80 - 0.86) for internal validation sets and 0.74 (95% CI: 0.67 – 0.89) for external validation sets. There was no statistically significant difference in the diagnostic performance between the two validation sets (p=0.42). Figure 2.B illustrates paired forest plots from the overall meta-analysis of these models, stratified by internal and external validation sets. The pooled sensitivity and specificity were as follows: for internal validation sets, 79.3% (95% CI: 75.7% - 82.6%) sensitivity and 77.2% (95% CI: 72.5% - 81.3%) specificity; for external validation sets, 71.9% (95% CI: 65.5% - 77.5%) sensitivity and 80.2% (95% CI: 54.6% - 93.2%) specificity.

**Figure 2.**
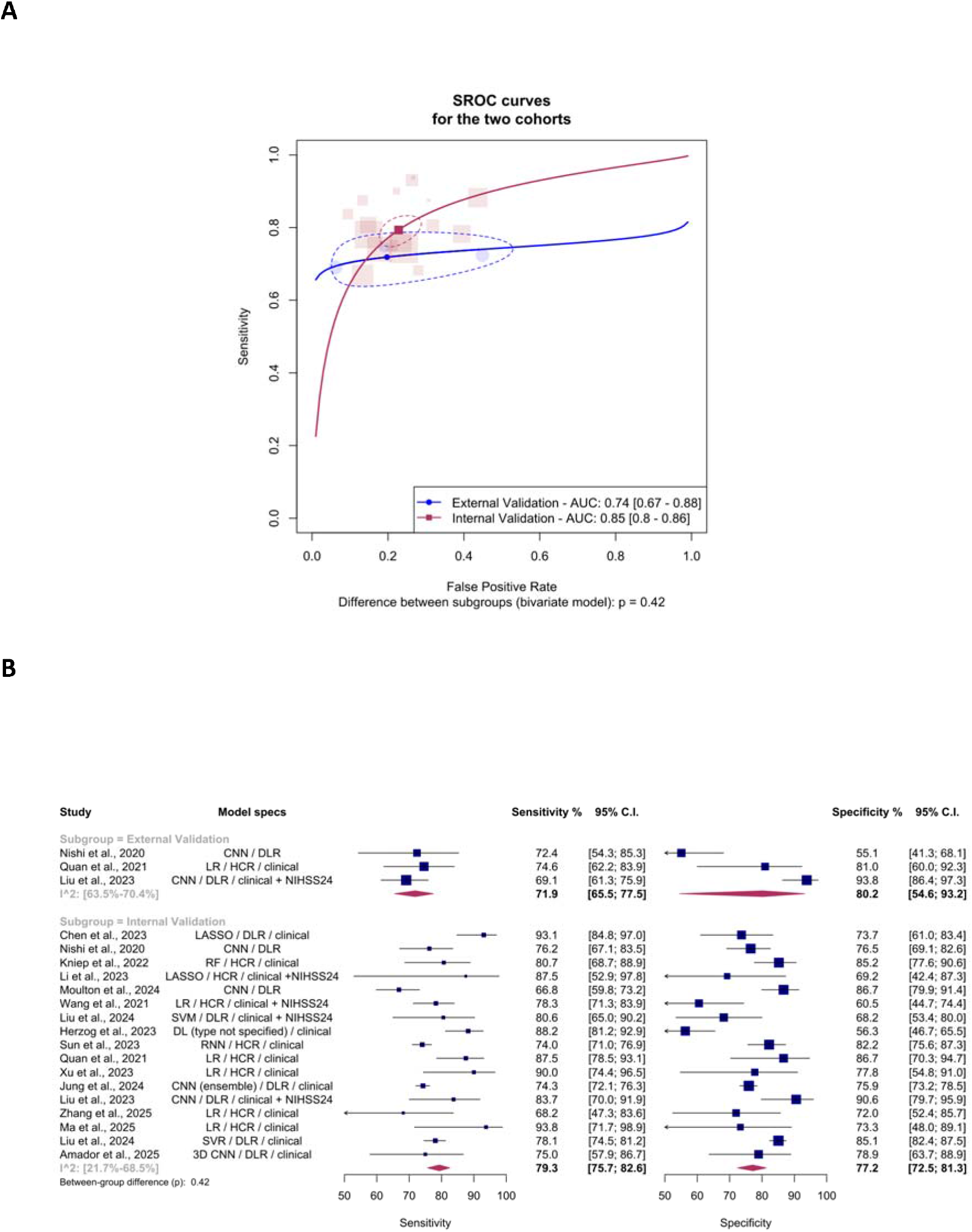
(A) Summary receiver-operating characteristic (SROC) curves with bivariate-model 95% confidence ellipses for models reported in external validation cohorts (AUC 0.74, 95% CI 0.67–0.88) and internal validation cohorts (AUC 0.85, 95% CI 0.80–0.86); between-subgroup difference p = 0.42. (B) Coupled forest plot of model-level sensitivity and specificity with subgroup summary diamonds (random-effects, bivariate model) and per-subgroup I² ranges.

Heterogeneity was observed across internal validation sets (I²: 21.7% - 68.5%) and external validation sets (I²: 63.5% - 70.4%). Leave-one-out analysis identified the study by Nishi et al., (2020) [33] as a significant outlier in the external validation sets. Revised forest plots, excluding this outlier, are presented in Figure E1. Even after excluding this outlier, no significant difference was found in diagnostic performance of the models in the two datasets (p=0.141).

### Subgroup analysis: deep learning vs. hand-crafted radiomics

Subgroup analyses were performed to compare models using DL algorithms with those based on HCR features in internal validation sets. Figure 3.A presents SROC curves for these models in internal validation sets, where the pooled AUC for DL models was 0.85 (95% CI: 0.77 – 0.88), compared to 0.86 (95% CI: 0.76 – 0.89) for HCR models. Figure 3.B shows the corresponding forest plots, where pooled sensitivity and specificity were as follows: for DL models, 77.3% (95% CI: 72.3% - 81.7%) sensitivity and 79.8% (95% CI: 74.9% - 83.9%) specificity; for HCR models, 79.4% (95% CI: 74.2% - 83.8%) sensitivity and 77.8% (95% CI: 70.8% - 83.5%) specificity. The difference between DL and HCR models was not statistically significant (p=0.667).

**Figure 3.**
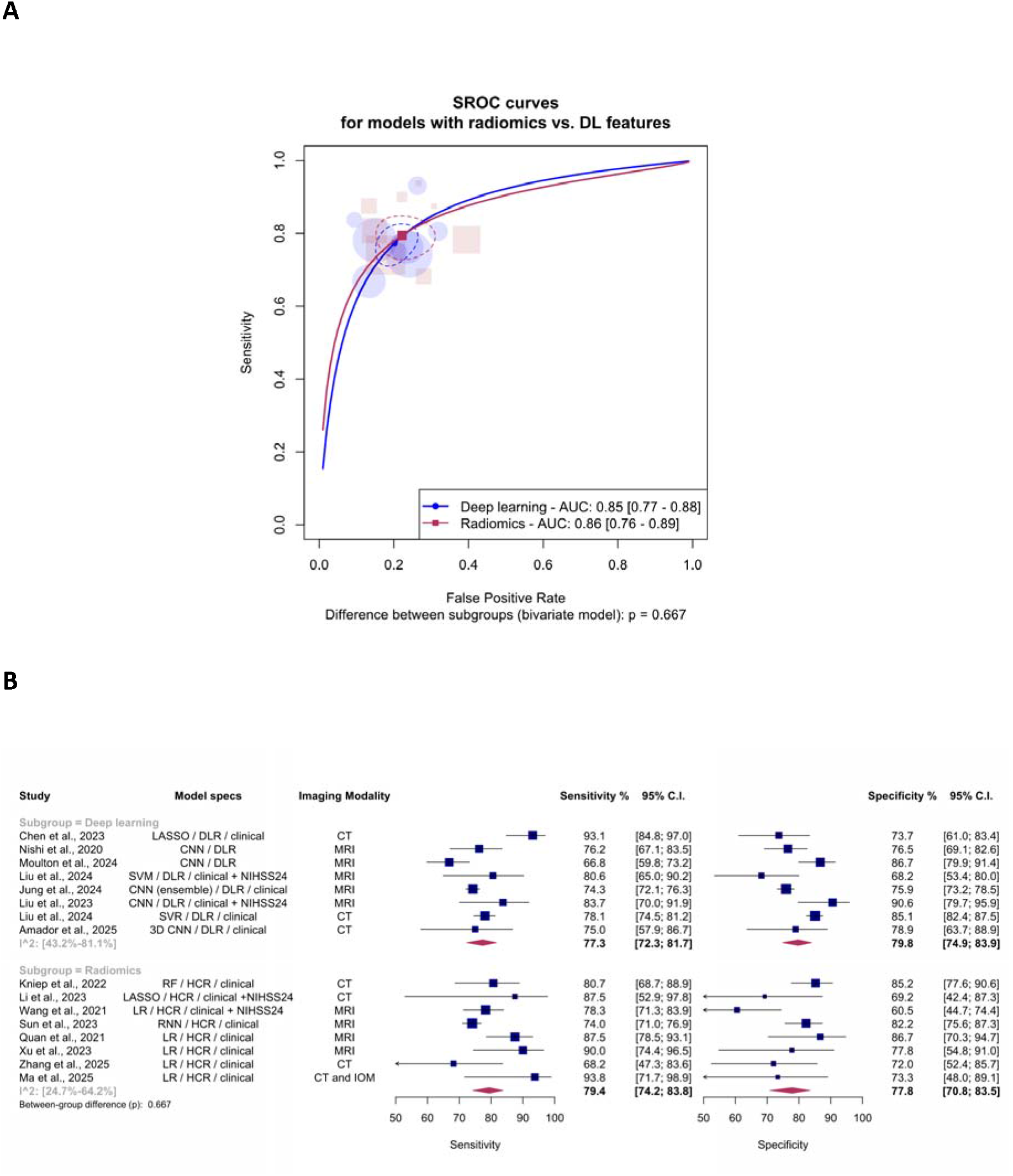
(A) SROC curves with 95% confidence ellipses comparing deep-learning–derived feature models (AUC 0.85, 95% CI 0.77–0.88) with handcrafted radiomic feature models (AUC 0.86, 95% CI 0.76–0.89); between-subgroup difference p = 0.667. (B) Coupled forest plot of sensitivity and specificity stratified by feature-extraction approach, with the imaging modality of each model indicated in the third column.

Heterogeneity was observed across DL models (I²: 43.2% - 81.1%) and HCR models (I²: 24.7% - 64.2%). Leave-one-out analysis identified the study by Wang et al., (2021) [36] as a significant outlier in the HCR subgroup. After excluding this study, the revised SROC curves and forest plots are shown in Figure E2 and Figure E3, respectively. After outlier exclusion, the differences between the subgroups became statistically significant (p=0.016); a post-hoc test indicated higher sensitivity for HCR than DL models (p=0.002). Given that this difference was contingent on outlier exclusion, it should be interpreted as exploratory.

### Subgroup analysis: comparing models with and without clinical data

A subgroup analysis was performed to compare models incorporating clinical data with those that did not, focusing on internal validation datasets. Figure 4.A displays the SROC curves, with the pooled AUC for models with clinical data being 0.86 (95% CI: 0.79 – 0.87) and 0.79 (95% CI: 0.75 – 0.82) for models without clinical variables (p< 0.01). Paired forest plots in Figure 4.B present pooled sensitivity and specificity as follows: for models incorporating clinical data, 80.6% (95% CI: 76.9% - 83.8%) sensitivity and 76.5% (95% CI: 71.2% - 81.1%) specificity; for models without clinical data, 71.2% (95% CI: 64.7% - 76.9%) sensitivity and 74.8% (95% CI: 69.0% - 79.9%) specificity. Significant differences were observed between these subgroups (p=0.007). Post-hoc analysis revealed that models with clinical features demonstrated significantly higher sensitivity compared to models without clinical features (p=0.004).

**Figure 4.**
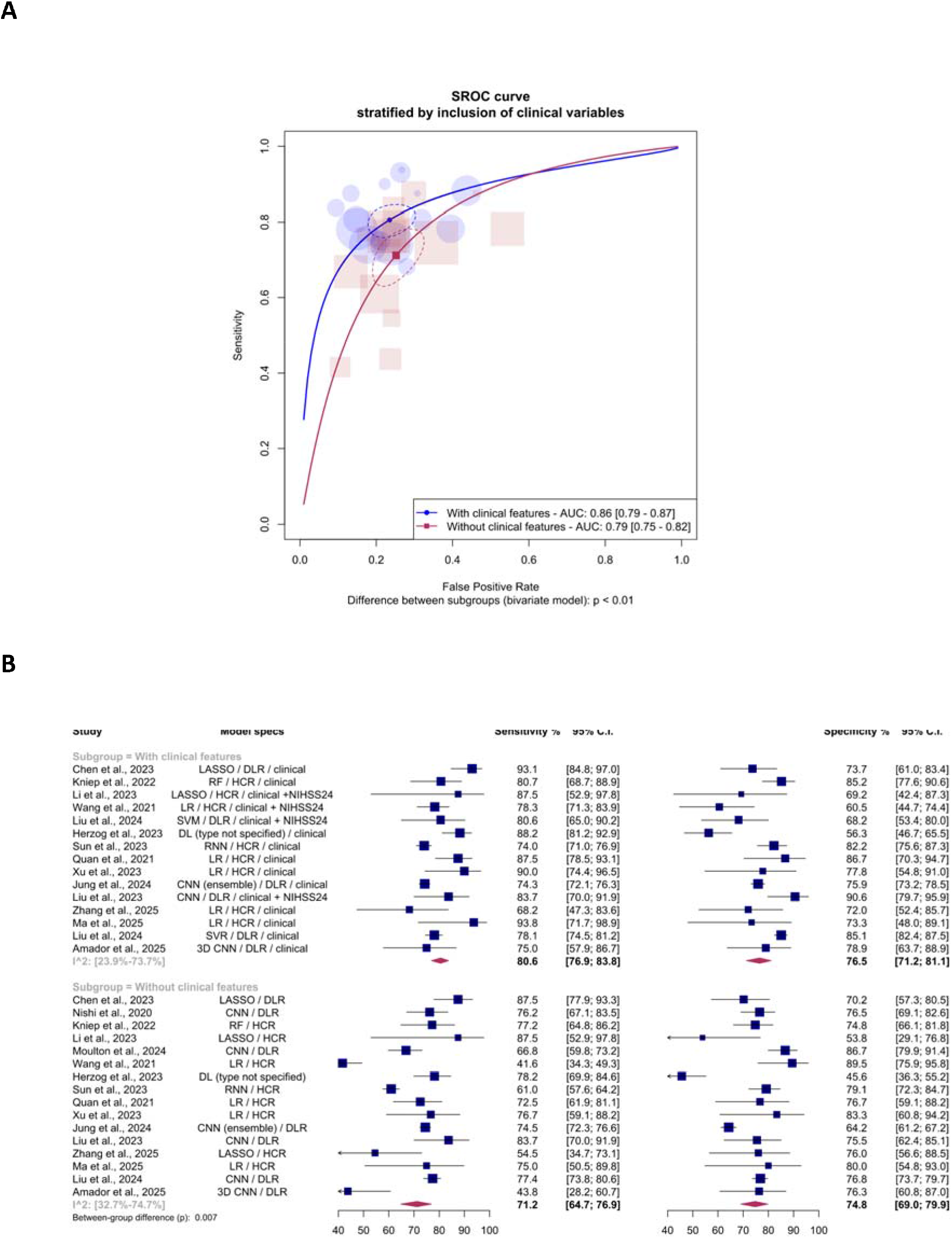
(A) SROC curves with 95% confidence ellipses for models that incorporated clinical features (AUC 0.86, 95% CI 0.79–0.87) versus those built on imaging-derived features alone (AUC 0.79, 95% CI 0.75–0.82); between-subgroup difference p < 0.01. (B) Coupled forest plot of sensitivity and specificity with bivariate subgroup summary diamonds (between-group p = 0.007).

Heterogeneity was observed across models with (I²: 23.9% - 73.7%) and without clinical features (I²: 32.7% - 74.7%). However, the leave-one-out analysis did not reveal any outliers explaining this heterogeneity.

### Subgroup analysis comparing models based on imaging modality (CT vs. MRI) among internal validation sets

In this subgroup analysis, models based on CT scans were compared with those based on MRI. As shown in Figure 5.A, the pooled AUC for CT-based models was 0.88 (95% CI: 0.8 – 0.9), compared to 0.83 (95% CI: 0.78 – 0.85) for MRI-based models. Paired forest plots in Figure 5.B show pooled sensitivity and specificity as follows: for CT-based models, 81.3% (95% CI: 76.5% - 85.3%) sensitivity and 81.2% (95% CI: 76.1% - 85.3%) specificity; for MRI-based models, 77.4% (95% CI: 73.2% - 81.0%) sensitivity and 74.9% (95% CI: 68.4% - 80.4%) specificity. The difference between the two modalities was not statistically significant (p=0.096).

**Figure 5.**
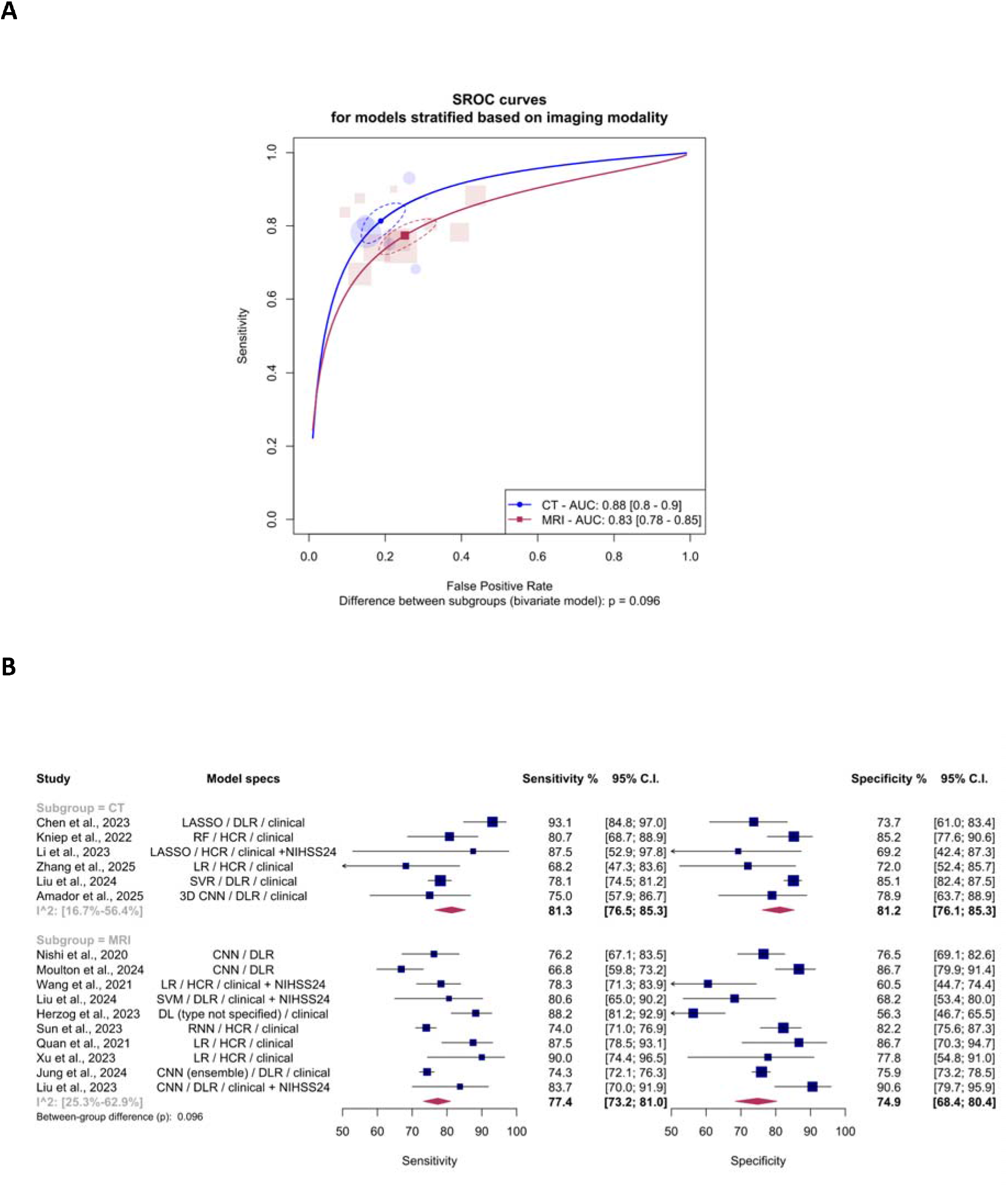
(A) SROC curves with 95% confidence ellipses for CT-based (AUC 0.88, 95% CI 0.80–0.90) and MRI-based (AUC 0.83, 95% CI 0.78–0.85) prediction models; between-subgroup difference p = 0.096. (B) Coupled forest plot of sensitivity and specificity stratified by imaging modality, with bivariate subgroup summary diamonds.

Heterogeneity was observed across MRI-based models (I²: 25.3% - 62.9%) and CT-based models (I²: 16.7% - 56.4%). Leave-one-out analysis identified the study by Zhang et al. (2025) [39] in the CT subgroup, and the studies by Liu et al. (2023) [30] and Quan et al. (2021) [34] as significant outliers in the MRI subgroup. After excluding these studies, the revised SROC curves and forest plots are shown in Figure E4 and Figure E5, respectively. According to the bivariate model, the differences between the subgroups became significant (p=0.002) after outlier exclusion. However, the post-hoc analysis did not reveal any significant differences regarding the two subgroups’ diagnostic performance. Because the global result was not corroborated by any pairwise contrast and depended on exclusion of three studies from already small subgroups, we did not interpret CT and MRI models as differing in diagnostic performance.

### Subgroup analysis comparing models based on treatment status (mechanical thrombectomy alone versus thrombolysis alone versus mechanical thrombectomy along with thrombolysis)

To assess the potential impact of treatment type on model performance, we conducted a subgroup analysis comparing models based on treatment status (MT alone vs. IVT alone vs. combined MT+IVT). Figure 6.A presents the SROC curves for this subgroup analysis in internal validation datasets. The pooled AUC for models using MT alone was 0.83 (95% CI: 0.68 – 0.87), for models using IVT alone was 0.93 (95% CI: 0.76 – 0.95), and for models using both MT and IVT was 0.87 (95% CI: 0.82 – 0.93) (p < 0.01).

**Figure 6.**
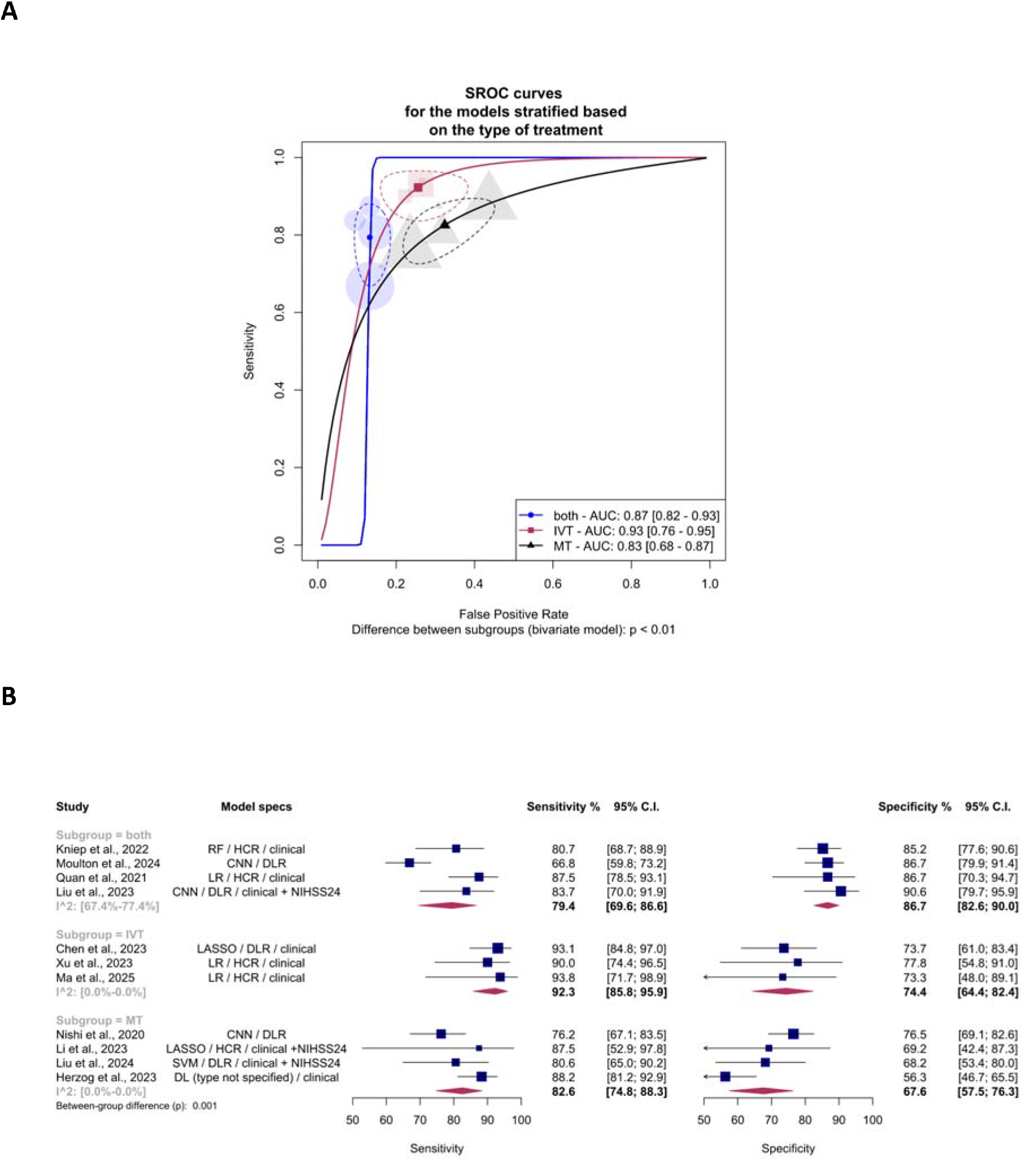
(A) SROC curves with 95% confidence ellipses for models trained on cohorts receiving both intravenous thrombolysis and mechanical thrombectomy (AUC 0.87, 95% CI 0.82–0.93), IVT alone (AUC 0.93, 95% CI 0.76–0.95), or MT alone (AUC 0.83, 95% CI 0.68–0.87); between-subgroup difference p < 0.01. (B) Coupled forest plot of sensitivity and specificity by treatment subgroup with bivariate summary diamonds (between-group p = 0.001).

Figure 6.B presents paired forest plots of sensitivity and specificity for each treatment group. The pooled sensitivity and specificity were as follows: for models predicting outcomes after MT alone, 82.6% (95% CI: 74.8% - 88.3%) sensitivity and 67.6% (95% CI: 57.5% - 76.3%) specificity; for models predicting outcomes following combined MT and IVT, 79.4% (95% CI: 69.6% - 86.6%) sensitivity and 86.7% (95% CI: 82.6% - 90.0%) specificity; for the IVT subgroup, 92.3% (95% CI: 85.8% - 95.9%) sensitivity and 74.4% (95% CI: 64.4% - 82.4%) specificity. The difference between subgroups was significant according to the bivariate model (p=0.001). Post-hoc analysis revealed that the models predicting outcomes following IVT alone had significantly higher sensitivity than the other groups; however, the wide AUC interval (0.93, 95% CI 0.76–0.95) indicates this estimate is imprecise and should be regarded as hypothesis-generating. Also the models predicting outcomes following combined MT+IVT had the highest specificity compared to the other groups (p< 0.05).

Heterogeneity was observed among models predicting outcomes following combined MT and IVT (I²: 67.4% - 77.4%). Leave-one-out analysis identified the study by Moulton et al., (2024) [32] as a significant outlier in this subgroup. After excluding this outlier study, the revised SROC curves and forest plots for this analysis are shown in Figure E6 and Figure E7. The difference between the subgroups remained significant after excluding the outlier (p=0.002).

### Publication bias

Figure 7 displays funnel plots visualizing the potential effect of publication bias in the overall meta-analysis of primary models from internal validation datasets. Based on Egger’s regression test, there was no evidence of publication bias (p= 0.11) in the internal validation sets.

**Figure 7.**
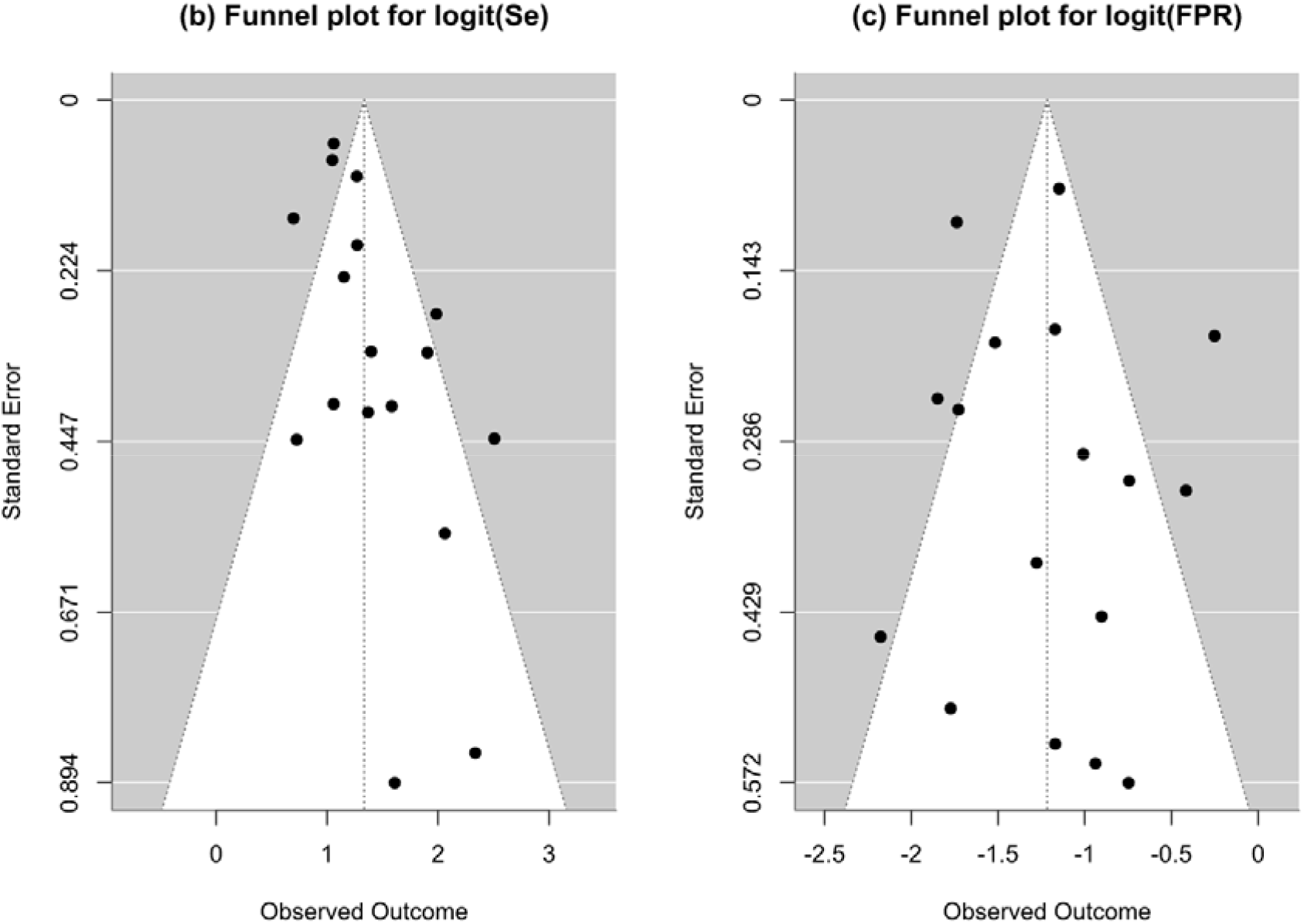
Publication bias assessment using funnel plot.

## Discussion

In this study, we assessed the role of DL and HCR models in predicting 90-day mRS score after ischemic stroke. The SROC curves revealed an AUC of 0.85 (95% CI: 0.80 - 0.86) for internal validation sets and 0.74 (95% CI: 0.67 - 0.89) for external validation sets, but it did not show a significant difference, suggesting a degree of generalizability in model performance across datasets. In addition, we conducted pre-specified sub-group analyses to determine the differences of different subgroups.

Firstly, regarding imaging modality, we observed that CT-based models were non-inferior to those based on MRI modalities, which predominantly included DWI. This finding is important as each modality varies in time requirements, accessibility, and the type of information it provides. NCCT and CTA are rapid and widely accessible imaging techniques essential for the initial evaluation of stroke patients. They are primarily used to exclude hemorrhage, detect early signs of ischemia, and identification of LVO [40]. CTP, on the other hand, offers advanced insights by measuring cerebral blood flow, blood volume, and time-to-peak parameters, which help in differentiating between salvageable brain tissue and irreversibly damaged areas [41]. On the other hand, DWI provides high-resolution images of the infarct core at very early stages, offering superior sensitivity compared to CT-based techniques [42]. Nevertheless, the higher costs, longer scan times, and limited accessibility of MRI machines, especially in emergency situations, make DWI less practical for immediate clinical decision-making compared to NCCT and CTA [43]. It is important to note that the performance of the models is highly heterogeneous and influenced by various factors, including acquisition parameters, pre-processing techniques, and model architecture.

Building on this, the non-inferior performance of CT-based models could have resulted from several factors. Firstly, CT scans are typically less prone to artifacts such as motion or susceptibility artifacts, which are more prevalent in DWI [44]. It should be noted that DL models can be especially sensitive to artifacts in medical imaging [45]. Additionally, the complexity inherent in DWI images necessitates larger sample sizes to train effective models without overfitting [46]. This explanation can also shed light on why some studies observed that the addition of PWI to DWI did not improve the performance [25, 47], as well as the results of another meta-analysis, which reported that CT radiomics was non-inferior to MRI for assessing plaque vulnerability [48].

Furthermore, regarding the integration of clinical data, which typically included parameters such as age and baseline NIHSS, with imaging features, our meta-analysis revealed a statistically significant improvement in model performance. This finding is consistent with other meta-analyses on radiomics models, which reported significant improvements in model performance after the inclusion of clinical variables [49, 50]. Notably, regarding the integration of clinical data with imaging features, as most of the studies adopted traditional statistical methods or conventional machine learning (ML) algorithms, there remains substantial room for improvement. Ensemble learning methods that combine the strengths of various ML algorithms with DL approaches can further enhance predictive performance [30].

Additionally, we did not observe a significant difference between HCR and DL models in predicting stroke outcomes. Among the DL models, most of the studies had applied ResNet architecture, alongside other architectures such as DeepSymNet and VGGNet, which differ in their structural design, such as the depth of layers, handling of feature extraction, and strategies for mitigating overfitting or vanishing gradients. The U-Net architecture was also utilized, notable for its design specifically tailored to automatic segmentation tasks, and therefore reducing bias [51]. Furthermore, emerging architectures like Transformer-based models are beginning to demonstrate promise in medical imaging by effectively capturing long-range dependencies and contextual information [52]. While most of the conducted studies have been on oncology [53–56], stroke prognosis prediction, too, might benefit from this innovation. Transformation-based data fusion has also been found to reduce the overfitting [24].

HCR, on the other hand, involves the extraction of a large number of quantitative features from medical images, aiming to capture the underlying tissue heterogeneity that may correlate with clinical outcomes [57]. For instance, texture features are commonly derived from methods like the gray-level co-occurrence matrix (GLCM), which quantifies spatial patterns and the intensity relationships between pixel pairs in an image [58]. Further, HCR can follow either manual or automated segmentation. Manual HCR entails expert delineation of regions of interest (ROIs) and the selective extraction of relevant features based on clinical insights. Notably, most of the HCR studies implemented manual segmentation, which often consisted of the stroke region. This approach, while allowing for tailored feature selection and potentially higher interpretability, is susceptible to inter-observer variability, which can affect reproducibility [59]. To address this limitation, incorporating automated segmentation methods in future studies could provide a more consistent and objective foundation, enhancing comparability with DL models and improving overall model performance evaluation.

As mentioned, conventional ML models based on HCR carry the benefit of being explainable [60]. Making DL models explainable is crucial for their adoption and trust in clinical settings, where understanding the rationale behind predictions can inform decision-making and ensure accountability. To achieve explainability, several strategies can be employed. Three studies implemented Grad-CAM (Gradient-weighted Class Activation Mapping) to visualize the regions of input images that most influence the model’s predictions [61–63].

Moreover, few studies assessed the predictive power of the models in posterior circulation strokes, which affect areas such as the brainstem, cerebellum, and occipital lobes. These strokes are inherently more challenging to image and analyze compared to anterior circulation strokes, and often have poor prognosis [64]. The intricate and compact anatomy of the posterior regions makes it difficult to detect and accurately delineate lesions using standard imaging modalities like NCCT and CTA [65], and these strokes frequently require DWI or perfusion imaging. Posterior circulation strokes are less common than anterior strokes, leading to fewer datasets available for training AI models. This data scarcity might impact the development and validation of predictive models. Encouraging multi-center collaborations to aggregate sufficient data for posterior circulation strokes could address this limitation and improve model generalizability.

Lastly, in this meta-analysis, we observed that models developed exclusively for patients undergoing MT exhibited lower sensitivity but comparatively higher specificity compared to those incorporating patients treated with both MT and IVT. This reduced sensitivity in MT-only models may be attributed to the fact that MT patients often present with larger infarctions, which have been shown to diminish the predictive power of models due to the increased variability and complexity associated with extensive brain damage [33]. Another contributing factor could be the variability in MT outcomes, which may depend significantly on the expertise of the thrombectomy operator and the specific techniques employed, further adding heterogeneity to the data. Conversely, the higher specificity suggests that when MT-only models do predict an unfavorable outcome, they do so with greater confidence, potentially reflecting clearer failure phenotypes in severe strokes.

## Limitations

This study has several limitations. First, there was notable heterogeneity among the included studies in baseline patient status, AI architectures, imaging protocols, and clinical variables incorporated into the models, so pooled estimates should be read as averages across methodologically distinct approaches. Second, several subgroup findings, the sensitivity advantage of handcrafted radiomics and the apparent CT–MRI difference, emerged only after exclusion of outlier studies and should be regarded as exploratory. Third, subgroups were small and confidence intervals correspondingly wide, particularly in the treatment-stratified analyses, and no correction was applied for multiple comparisons. Fourth, external validation was reported in a minority of studies, so the comparable internal and external performance observed may reflect limited external evidence rather than genuine transportability. Finally, most studies focused on anterior circulation strokes and few assessed model explainability. Future research should address these gaps through standardized protocols, independent external validation, and greater attention to explainable models and underrepresented stroke subtypes.

## Conclusion

In conclusion, this study shows the promising potential of DL and HCR models based on imaging modalities including CT and MRI for predicting stroke outcomes. Notably, models integrating imaging with routinely available clinical variables such as age and baseline NIHSS outperformed imaging-only models, indicating that imaging-derived features are best deployed as a complement to, rather than a substitute for, established clinical predictors. Future research should focus on improving these models by incorporating newer DL architectures, such as transformers, while HCR models can benefit from automated segmentation.

## Data Availability

Not applicable as this is a review paper.

## Declarations

- Ethics and Consent to Participate declarations: not applicable.
- Funding information: This study was not funded.
- Conflict of interest: Authors declare no conflict of interest.

