## Supplementary Material for "Deep Learning and Radiomics Models for Predicting 90-Day Outcomes in Ischemic Stroke: A Systematic Review and Meta-Analysis"

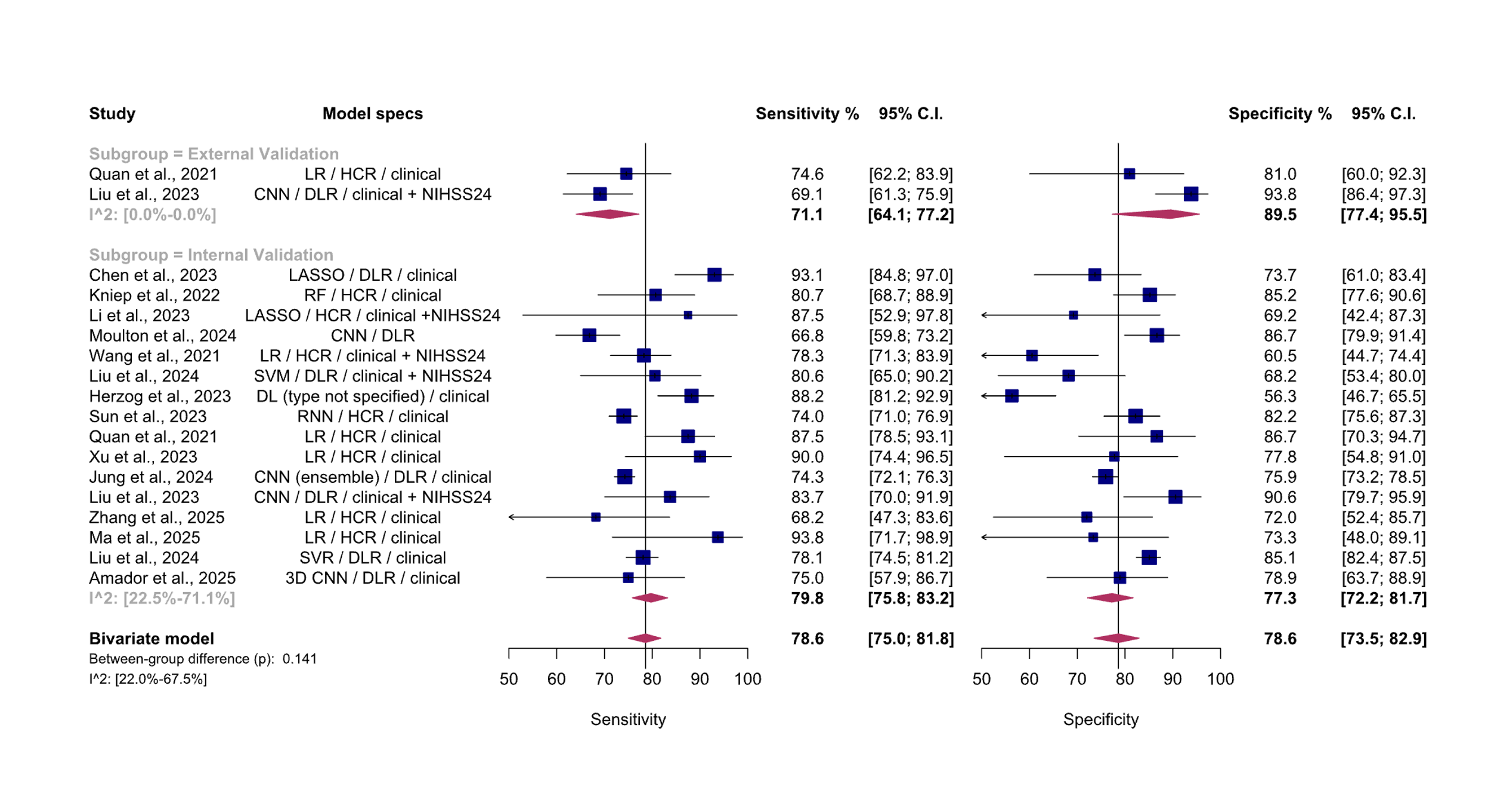
 **Figure E1.** Paired forest plots of the general meta-analysis of the main models of the included studies, stratified based on internal and external validation sets; after omitting one outlier study. CI: Confidence interval. CNN: Convolutional neural network. DL: Deep learning. DLR: Deep learning radiomics. HCR: handcrafted radiomics. LASSO: Least absolute shrinkage and selection operator. LR: Logistic regression. NIHSS24: National Institutes of Health Stroke Scale measured at 24 hours. RF: Random forest. SVM: Support vector machine. The between-group difference is derived from the bivariate model.

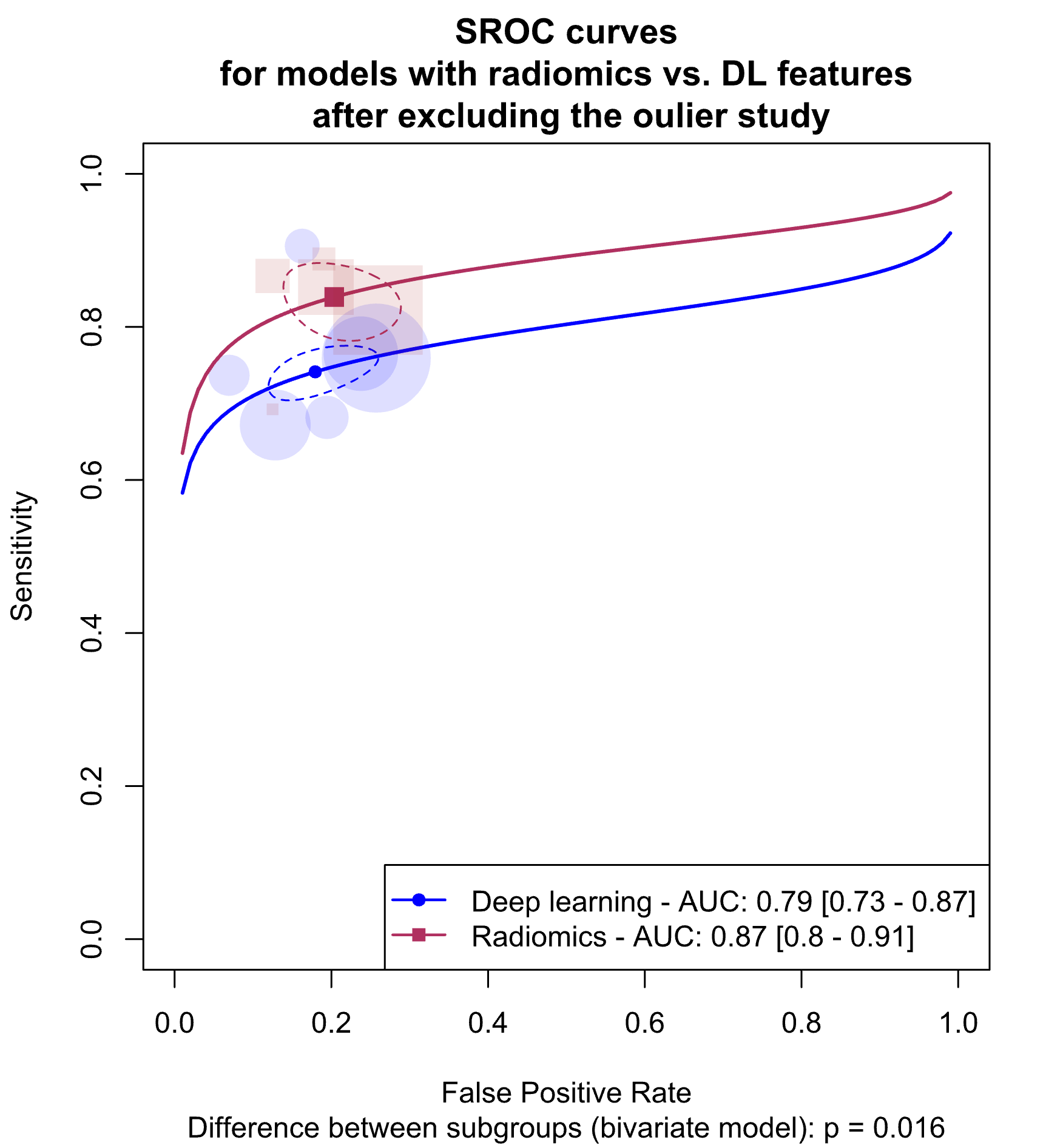

**Figure E2.** Summary Receiver Operating Curves (SROCs) for subgroup meta-analysis comparing deep learning and hand-crafted radiomics models in internal validation sets., after removing the outlier study. AUC: Area Under the Curve. DL: Deep learning. The between-group difference is derived from the bivariate model.

**Figure E3.** Paired forest plots for the subgroup meta-analysis comparing deep learning and hand-crafted radiomics models in internal validation sets, after excluding one outlier study. CI: Confidence interval. CNN: Convolutional neural network. DL: Deep learning. DLR: Deep learning radiomics. HCR: handcrafted radiomics. LASSO: Least absolute shrinkage and selection operator. LR: Logistic regression. NIHSS24: National Institutes of Health Stroke Scale measured at 24 hours. RF: Random forest. SVM: Support vector machine. The between-group difference is derived from the bivariate model.

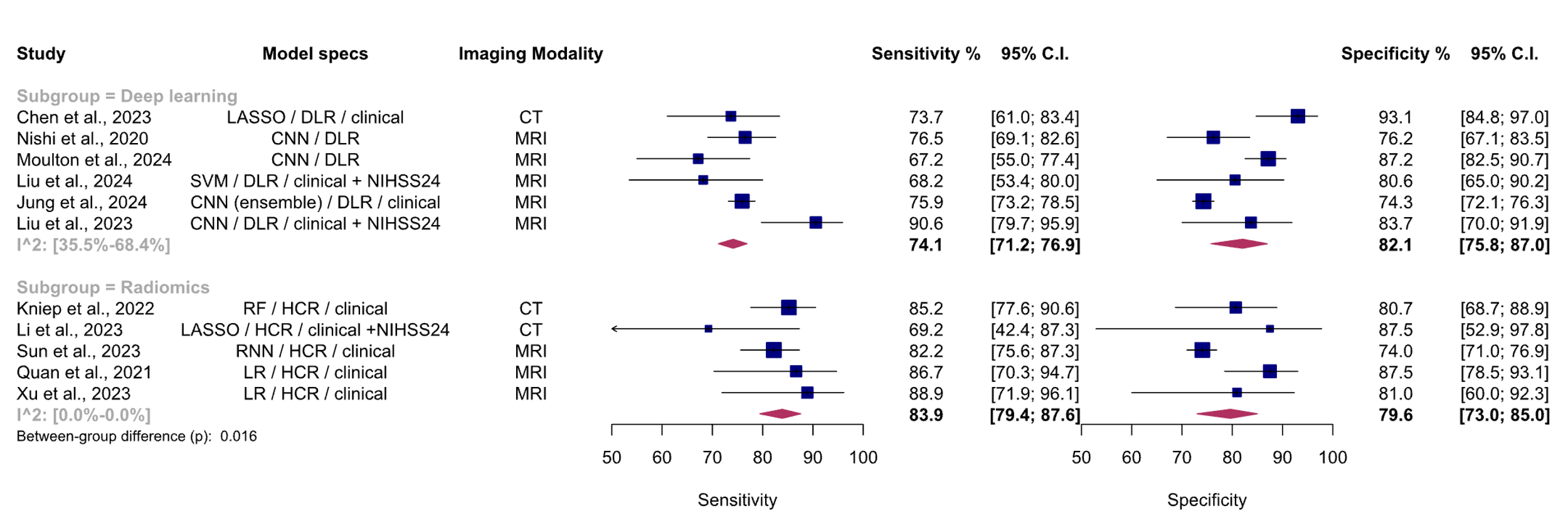

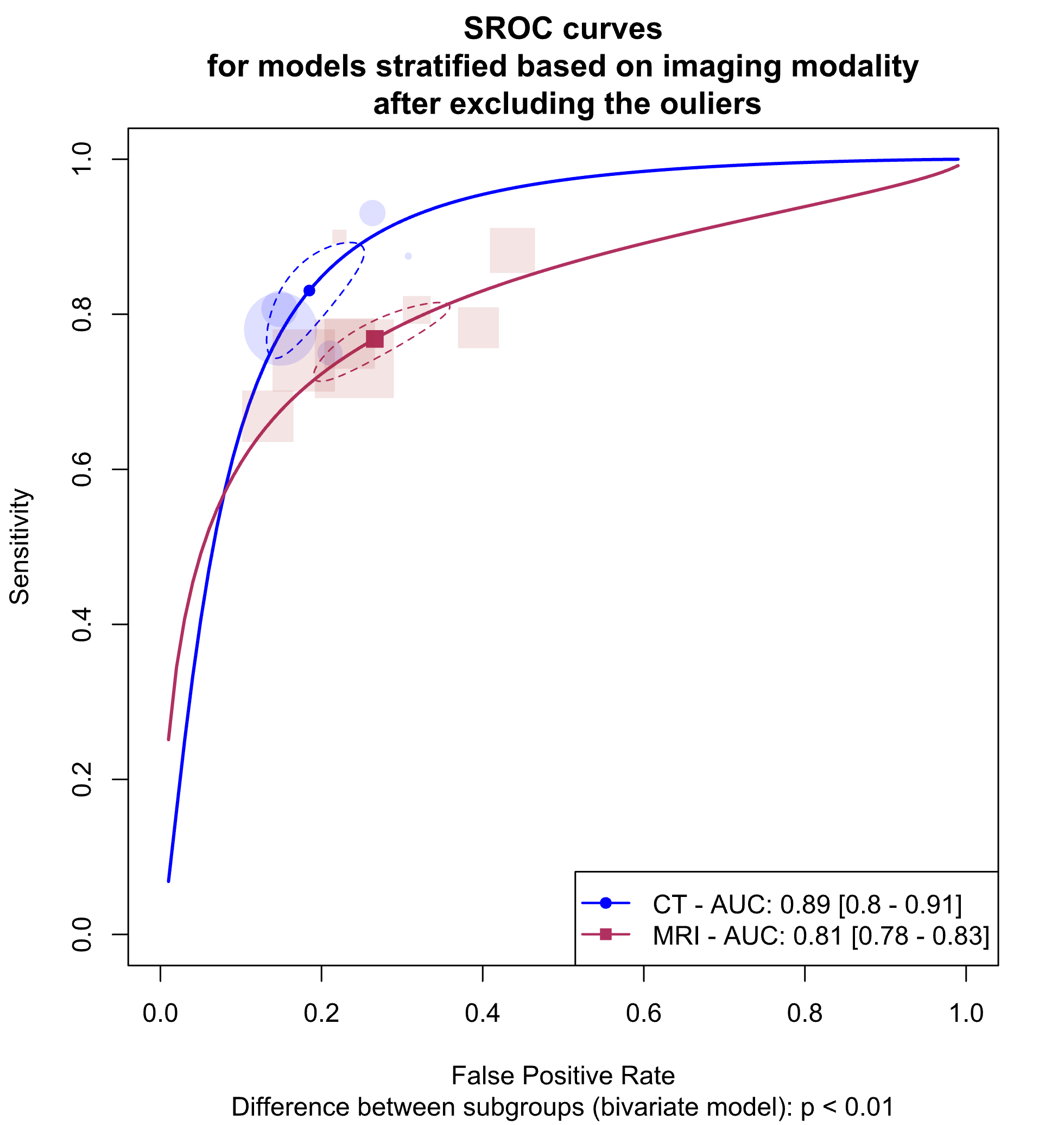

**Figure E4.** Summary Receiver Operating Curves (SROCs) for subgroup meta-analysis comparing models based on CT scan and MRI in internal validation sets, after excluding one outlier study. AUC: Area Under the Curve. The between-group difference is derived from the bivariate model.

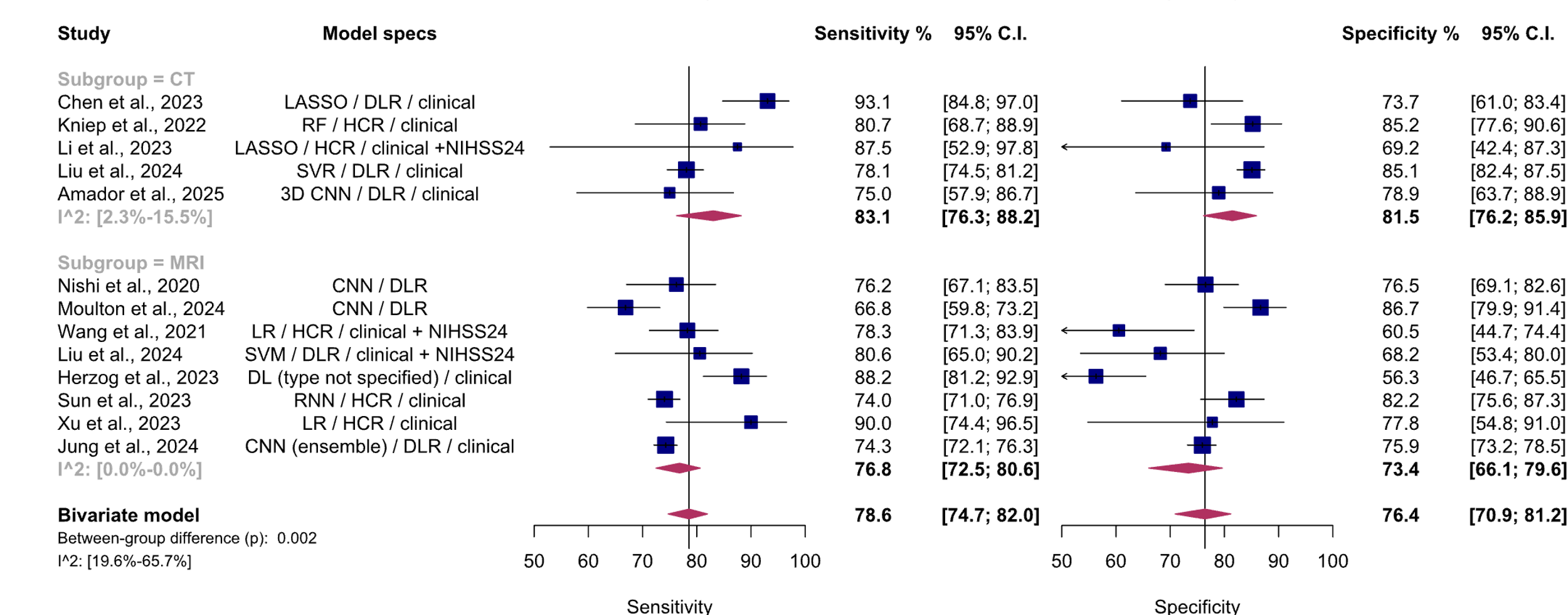

**Figure E5.** Paired forest plots of the subgroup meta-analysis for subgroup meta-analysis comparing models based on CT scan and MRI in internal validation sets, after excluding one outlier study. CI: Confidence interval. CNN: Convolutional neural network. DL: Deep learning. DLR: Deep learning radiomics. HCR: handcrafted radiomics. LASSO: Least absolute shrinkage and selection operator. LR: Logistic regression. NIHSS24: National Institutes of Health Stroke Scale measured at 24 hours. RF: Random forest. SVM: Support vector machine. Between group difference is derived from bivariate model.

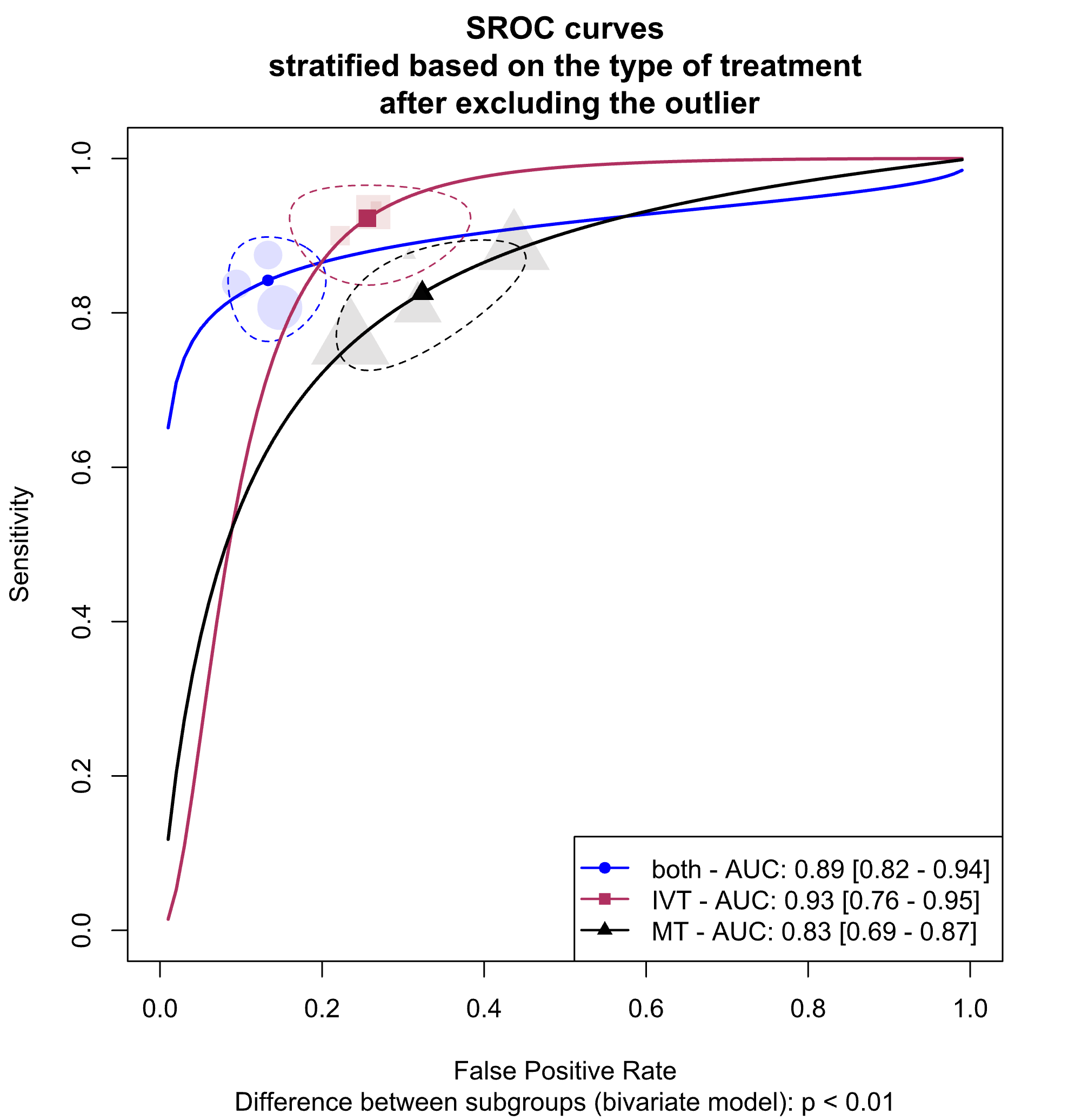

**Figure E6.** Summary Receiver Operating Curves (SROCs) for subgroup meta-analysis comparing models based the type of treatment (mechanical thrombectomy alone vs. mechanical thrombectomy+ thrombolysis) in internal validation sets, after excluding one outlier study. AUC: Area Under the Curve. MT: Mechanical thrombectomy. The between-group difference is derived from the bivariate model.

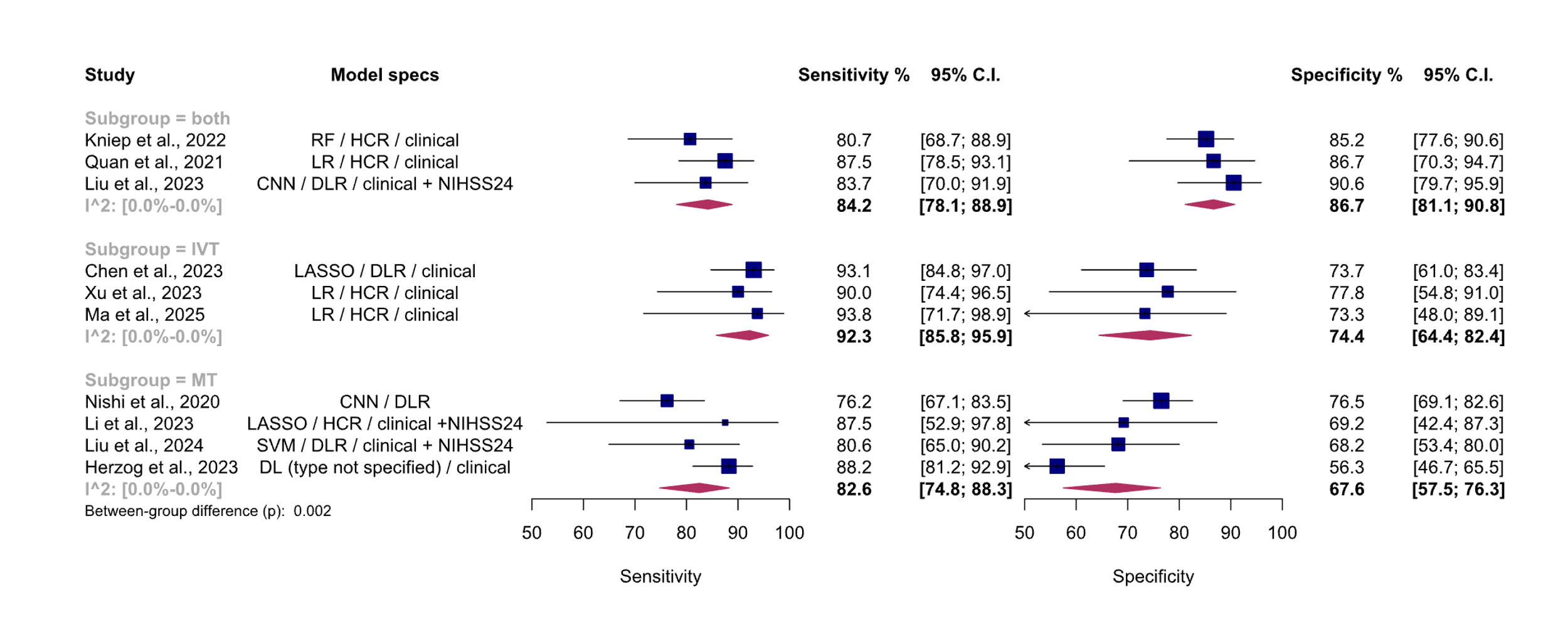

**Figure E7.** Paired forest plots of the subgroup meta-analysis for subgroup meta-analysis comparing models based on the type of treatment in internal validation sets, after excluding one outlier study. CI: Confidence interval. CNN: Convolutional neural network. DL: Deep learning. DLR: Deep learning radiomics. HCR: handcrafted radiomics. IVT: IV thrombolysis. LASSO: Least absolute shrinkage and selection operator. LR: Logistic regression. MT: Mechanical thrombectomy. NIHSS24: National Institutes of Health Stroke Scale measured at 24 hours. RF: Random forest. SVM: Support vector machine. Between group difference is derived from bivariate model.

**Table E1.** Included search terms.

| Searched database | Search syntax |
| --- | --- |
| PubMed, Scopus, Web of Science and Embase | (stroke OR "cerebral infarction" OR "brain infarction") AND (mrs OR "modified Rankin Scale") AND (radiomics OR HCR OR "deep learning" OR "convolutional network*" OR "neural network*" OR CNN OR GAN OR "Generative adversarial network*") AND (ct OR "computed tomography" OR MRI OR DWI OR “diffusion weighted imaging” OR DTI OR “diffusion tensor imaging” OR ADC OR “apparent diffusion coefficient”) |

**Table E3. Quality assessment of included studies**
